# Attitudes toward COVID-19 illness and COVID-19 vaccination among pregnant women: a cross-sectional multicenter study during August-December 2020

**DOI:** 10.1101/2021.03.26.21254402

**Authors:** Ashley N. Battarbee, Melissa S. Stockwell, Michael Varner, Gabriella Newes-Adeyi, Michael Daugherty, Cynthia Gyamfi-Bannerman, Alan Tita, Kelly Vorwaller, Celibell Vargas, Akila Subramaniam, Lawrence Reichle, Romeo Galang, Emily Powers, Miriam Lucca-Susana, Mickey Parks, Tiffany J. Chen, Hilda Razzaghi, Fatimah S. Dawood

## Abstract

**Objective:** Evaluate pregnant women’s attitudes toward COVID-19 illness and vaccination and identify factors associated with vaccine acceptability.

**Study Design:** Cross-sectional survey among pregnant women enrolled in a prospective COVID-19 cohort study in Salt Lake City, UT, Birmingham, AL, and New York, NY, August 9– December 10, 2020. Women were eligible if they were 18-50 years old and <28 weeks of gestation. Upon enrollment, women completed surveys regarding concerns about COVID-19 illness and likelihood of getting COVID-19 vaccine if one were available during pregnancy. Vaccine acceptability was defined as a response of “very likely” or “somewhat likely” on a 4-point Likert scale. Factors associated with vaccine acceptability were assessed with multivariable logistic regression.

**Results:** Of 939 pregnant women eligible for the main cohort study, 915 (97%) consented to participate. Among these 915 women, 39% self-identified as White, 23% Black, 33% Hispanic, and 4% Other. Sixty-two percent received an influenza vaccine last season. Seventy-two percent worried about getting sick with COVID-19. If they were to get sick, 92% worried about harm to their pregnancy and 80% about harm to themselves. Only 41% reported they would get a vaccine. Of women who were unlikely to get vaccinated, the most frequently cited concern was vaccine safety for their pregnancy (82%). Non-Hispanic Black and Hispanic women had lower odds of accepting a vaccine compared with non-Hispanic White women (adjusted odds ratios (aOR) 0.4, 95%CI 0.2–0.6 for both). Receipt of influenza vaccine during the previous season was associated with higher odds of vaccine acceptability (aOR 2.1, 95%CI 1.5-3.0).

**Conclusion:** Although most pregnant women worried about COVID-19 illness, <50% were willing to get vaccinated during pregnancy. Racial and ethnic disparities in plans to accept COVID-19 vaccine highlight the need to prioritize strategies to address perceived barriers among groups at high risk for COVID-19.

## INTRODUCTION

As of March 1, 2021, there have been over 114 million cases of COVID-19 with more than 2.5 million deaths worldwide(1). Of the 1.1 million cases in the United States, there have been more than 73,000 laboratory-confirmed cases of COVID-19 among pregnant women(2). Based on accumulating data, pregnant women appear to be at increased risk for severe COVID-19 disease(3, 4). In a recent report that included 400,000 women of reproductive age, pregnant women with COVID-19 were found to be more likely than non-pregnant women to be admitted to the intensive care unit, receive extracorporeal membrane oxygenation, and die(5). Comparisons of rates of SARS-CoV-2 infection and risks for COVID-19 disease among pregnant versus non-pregnant women in the United States also suggest that Hispanic women may be at increased risk of infection and Hispanic and non-Hispanic Black women may be at increased risk of severe disease(5, 6). At this time, the risk of transplacental transmission of COVID-19 to the developing fetus appears to be low,(7, 8) and most studies have shown no increased risk of spontaneous abortions or stillbirth(9, 10). However, pregnant women with COVID-19 illness may be at increased risk for preterm birth(11).

Although social distancing, hand hygiene, and mask wearing are key non-pharmaceutical interventions to slow the spread of COVID-19, widespread safe and effective vaccination is ultimately necessary to control this global pandemic(12). Many COVID-19 vaccine trials are currently being conducted worldwide with many vaccines in Phase III testing and now a few COVID-19 vaccines approved for emergency use (14, 15). Pregnant women have not been included in the Phase III trials. However, both the American College of Obstetricians and Gynecologists (ACOG) and the Society for Maternal Fetal Medicine (SMFM) recommend that pregnant women be included in vaccine trials and offered COVID-19 vaccines (16, 17). In addition, the Centers for Disease Control and Prevention recommends that pregnant women be provided the opportunity to choose whether to receive COVID-19 vaccine under the current Emergency Use Authorization(18, 19). However, the willingness of pregnant women to be vaccinated is unknown.

We evaluated pregnant women’s attitudes toward COVID-19 illness and vaccination during pregnancy and assessed factors associated with vaccine acceptability. Based on findings from studies of influenza vaccine acceptance among adults,(20) we hypothesized that women from racial and ethnic minority groups would be less willing to be vaccinated.

## METHODS

We performed a cross-sectional study of pregnant women enrolled August 9– December 10, 2020, in the Epidemiology of Severe Acute Respiratory Syndrome in Pregnancy and Infancy (ESPI) Community Cohort, an ongoing prospective longitudinal cohort study conducted at three centers in the United States (Birmingham, AL; Salt Lake City, UT; and New York, NY). The ESPI Community Cohort study is designed to estimate the incidence of SARS-CoV-2 infection, identify risk factors for infection, and characterize the clinical spectrum of infection among pregnant women receiving prenatal care at the three study sites. Centralized Institutional Review Board approval was obtained (IRB-AAAT1906), and informed consent was obtained from all participants.

Women were eligible for participation in the main cohort if they were 18–50 years old and were at less than 28 weeks of gestation to allow for an average of at least 12 weeks of surveillance time in the cohort prior to end of pregnancy. Women without a functioning telephone and those who were not willing to respond to weekly COVID-19 surveillance questionnaires or to self-collect nasal swabs weekly were not eligible for participation, as these were key components of the ESPI Community Cohort study. Additionally, women who were unable to speak and read either English or Spanish, and those currently enrolled in a COVID-19 or influenza vaccine trial, were not eligible.

Women were not excluded based on prior suspected or confirmed COVID-19 infection. Study staff attempted to approach all women who met age and gestational age criteria for enrollment among those receiving prenatal care at the three participating centers during the study period. Participants were recruited over the phone and in person at outpatient prenatal care offices. Women who consented to participate in the ESPI Community Cohort were asked to complete a standardized survey at the time of study enrollment. The survey included questions about maternal demographics and socioeconomic characteristics, past medical and obstetric history, and attitudes towards SARS-CoV-2 infection/COVID-19 illness and vaccination in pregnancy. Women were considered fully enrolled in the main ESPI Community Cohort study if they consented to participation and completed eight core questions on the enrollment survey about enrollment date, estimated delivery date, number of gestations, diagnosis of gestational hypertension or gestational diabetes during the current pregnancy, presence of underlying medical conditions, and number of prior pregnancies.

Using a 4-point Likert scale, pregnant women were asked “How worried are you about getting sick with COVID-19?” as well as “If you were to get sick with COVID-19, how worried are you that COVID-19 would harm <u>you</u>?” and “…how worried are you that COVID-19 would harm <u>your pregnancy</u>?” Women were deemed concerned if they answered “very worried” or “somewhat worried.” Women were asked about their most trusted source for receiving information about COVID-19, and women were also asked “If a COVID-19 vaccine were to become available for pregnant women, how likely would you be to get the vaccine for yourself during your pregnancy?” Vaccine acceptability was defined as women who answered “very likely” or “somewhat likely.” Women were considered not willing to get a vaccine if they answered “not too likely” or “not at all likely.” Lastly, women were asked to answer multiple choice questions about the reasons why they would (or would not) get a vaccine; in addition to standard response choices, questions included an option to indicate other reasons with free text entry. Answer choices for reasons for getting the vaccine included: “to protect myself from getting sick with COVID-19”, “to protect my pregnancy,” “to protect others in my family,” and “to protect others in the community from getting sick with COVID-19.” Answer choices for reasons for not getting the vaccine included: “concerns or question about vaccine safety for myself,” “concerns or questions about vaccine safety for my pregnancy,” “concerns or questions about whether the vaccine would work to protect me from COVID-19,” and “I don’t think I need the vaccine.”

A complete case analysis was performed including women who enrolled in the cohort study from August 9 to December 10, 2020. Baseline maternal demographic and obstetric characteristics in the population were described in aggregate and by vaccine acceptability. Descriptive statistics were used to summarize pregnant women’s responses to questions about COVID-19 illness and vaccination during pregnancy. Logistic regression was used to estimate the association between baseline characteristics and vaccine acceptability. Key characteristics assessed included study site, maternal age, self-reported race and ethnicity, education, employment status, employment in a healthcare occupation, household income, maternal medical comorbidities, and prior receipt of influenza vaccination during the 2019–20 influenza season. Race and ethnicity were characterized as Black if women identified as Non-Hispanic Black or African American, White if women identified as Non-Hispanic White, and Hispanic if women identified as Hispanic or Latino regardless of race. All characteristics that were significantly associated with vaccine acceptability in unadjusted analyses at p<0.20 were included in the multivariable logistic regression model, except for study site and employment status, which were highly correlated with other covariates. Backward step-wise selection using a p-value cut-off of 0.2 and evaluating for a >10% change in adjusted odds ratios was used to achieve a final, parsimonious model from which adjusted odds ratios and 95% confidence intervals (CI) were estimated. To assess for differences in predictors of vaccine acceptability before and after the first release of phase III COVID-19 vaccine trial results in the United States in early November 2020, multivariable models were run among all women and stratified by women who completed enrollment surveys during August 9-November 14 and November 15-December 20, 2020 using the same explanatory variables selected for the final primary model. Adjusted odds ratios and 95% confidence intervals (CI) were estimated.

The associations between: (1) women’s level of concern about getting sick with COVID-19 and (2) women’s level of concern about harm to themselves or their pregnancy from COVID-19 in the setting of SARS-CoV-2 infection and vaccine acceptance were also assessed using bivariate analysis.

SAS Version 9.4 was used for statistical analysis, and statistical significance was set at p=0.05.

## RESULTS

During the study period, 1186 women were screened for eligibility for the main cohort study, of whom 939 (79%) were eligible. Among these 939 women, 915 (97%) consented to participate in the main cohort study and were fully enrolled.

Among the 915 women included in this analysis, 39% self-identified as White, 23% as Black, and 33% as Hispanic. Overall, 64% of women had more than a high school education, 60% were employed, and 20% lived in households with income below the local poverty line. Twenty-eight percent of women had one or more underlying medical conditions. Last season 62% of women reported receiving the influenza vaccine (Table 1).

**Table 1.** Baseline demographic and other characteristics (n=915)

| Table 1. Baseline demographic and other characteristics (n=915) |  |  |  |  |
| --- | --- | --- | --- | --- |
|  | Total<br>N=915<br>n (col %) | Willing to get<br>vaccine*<br>n=374<br>n (row %) | Not willing to<br>get vaccine*<br>n=429<br>n (row %) | p-value |
| Site |  |  |  |  |
| Birmingham, AL | 267 (29) | 98/245 (40) | 147/245 (60) | <0.01 |
| Salt Lake City, UT | 340 (37) | 196/311 (63) | 115/311 (37) |  |
| New York City, NY | 308 (34) | 80/247 (32) | 167/247 (68) |  |
| Age, years |  |  |  |  |
| 18-34 | 741 (81) | 295/647 (46) | 352/647 (54) | 0.22 |
| 35-50 | 172 (19) | 79/155 (51) | 76/155 (49) |  |
| Race/ethnicity |  |  |  |  |
| White, non-Hispanic | 346 (39) | 202/319 (63) | 117/319 (37) | <0.01 |
| Black, non-Hispanic | 201 (23) | 58/186 (31) | 128/186 (69) |  |
| Hispanic | 293 (33) | 83/238 (35) | 155/238 (65) |  |
| Other, non-Hispanic | 38 (4) | 16/30 (53) | 14/30 (47) |  |
| Education |  |  |  |  |
| Less than high school diploma | 57 (6) | 23/52 (44) | 29/52 (56) | <0.01 |
| High school diploma | 265 (29) | 89/225 (40) | 136/225 (60) |  |
| Some college/technical school | 230 (25) | 63/191 (33) | 128/191 (67) |  |
| College degree | 214 (23) | 106/193 (55) | 87/193 (45) |  |
| Graduate degree | 147 (16) | 93/141 (66) | 48/141 (34) |  |
| Household below poverty level* | 177 (20) | 73/161 (45) | 88/161 (55) | 0.71 |
| Marital status |  |  |  |  |
| Married | 473 (52) | 235/425 (55) | 190/425 (45) | <0.01 |
| Never married | 267 (30) | 83/224 (37) | 141/224 (63) |  |
| Committed partnership | 127 (14) | 39/112 (35) | 73/112 (65) |  |
| Other | 37 (4) | 15/33 (45) | 18/33 (55) |  |
| Employed | 549 (60) | 239/493 (48) | 254/493 (52) | 0.23 |
| Occupation<sup>†</sup> |  |  |  |  |
| Healthcare professional | 228 (42) | 108/203 (53) | 95/203 (47) | 0.10 |
| Other | 316 (58) | 130/285 (46) | 155/285 (54) |  |
| ≥1 underlying medical condition<sup>‡</sup> | 253 (28) | 107/232 (46) | 125/232 (54) | 0.87 |
| Previously had<br>suspected/confirmed COVID-19<br>before enrollment | 44 (5) | 20/37 (54) | 17/37 (46) | 0.64 |
| Received Influenza Vaccine Last<br>Year | 564 (62) | 273/497 (55) | 224/497 (45) | <0.01 |
| *Based on number of household members: https://aspe.hhs.gov/poverty-guidelines.<br>Column % do not include missing.<br>If data are missing for a variable, n included are indicated in parentheses after variable.<br>*Willing to get vaccine defined as survey response of “very likely” or “somewhat likely”. Not willing to get vaccine defined as “not likely” or “not at all likely.” Women who responded “unknown” are not included in the analysis (n=103).<br>†Among all employed (n=549)<br>‡Defined as asthma, chronic lung, metabolic, hematologic, cardiovascular, renal, hepatic, neurologic, rheumatologic disease or hypertension.<br>Total Missing/Decline to respond/Unknown: age (2, <1%), race/ethnicity (37, 4%), education (2, <1%), households below poverty level (15, 2%), marital status (11, 1%), employed (7, <1%), occupation (5, <1%), underlying conditions (1, <1%), previous COVID-19 (2, <1%), received influenza vaccine last year (21, 2%).<br>Abbreviations: IQR, interquartile range. |  |  |  |  |

Seventy-two percent (95% CI: 69%-75%) of women stated they were worried about getting sick with COVID-19. If they were to get sick with COVID-19, 92% (95% CI: 91%-94%) of women were worried that COVID-19 would harm their pregnancies, and 80% (95% CI: 77%-82%) were worried that COVID-19 would harm them (Figure 1). When asked what source of COVID-19 information women trusted the most, the most common answer was their obstetrician/gynecologist (42%, 95% CI: 38%-45%), followed by their family doctor or primary care provider (28%, 95% CI: 25%-30%), CDC (13%, 95% CI: 11%-15%), and other medical professionals (4%, 95% CI: 3%-5%).

**Figure 1.**
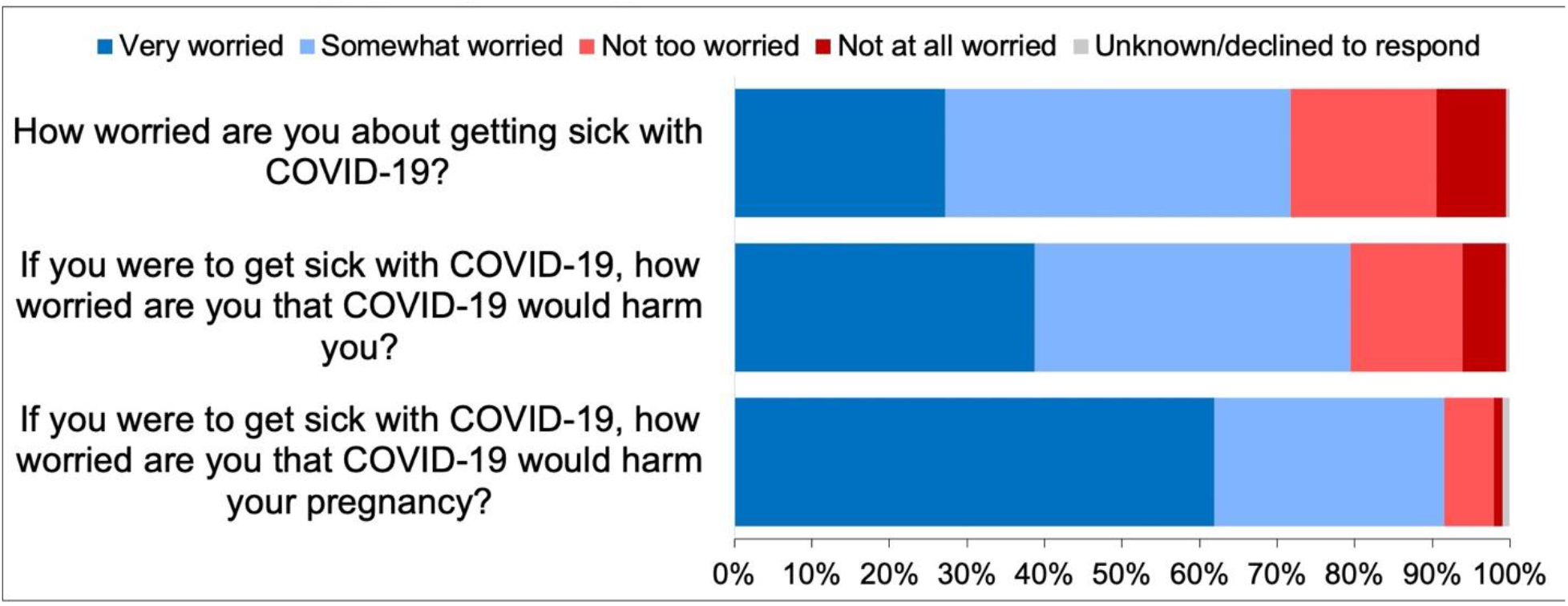
**Pregnant women’s attitudes towards COVID-19 illness during pregnancy, N=915**

Overall, 41% (374/915, 95% CI: 38%-44%) of women reported they would get a COVID-19 vaccine if one became available during their pregnancy. When stratified by enrollment month, the proportion of women willing to get a COVID-19 vaccine was similar (data not shown). Among women who were willing to get a vaccine during pregnancy, the most commonly cited reason for vaccine acceptability was to protect their pregnancy (95%, 95% CI: 93%-98%). Other reasons included protecting themselves (85%, 95% CI: 81%-89%), protecting family members (79%, 95% CI: 75%-83%), and protecting the community (68%, 95% CI: 63%-73%). In contrast, women who were not willing to get a vaccine during pregnancy most frequently cited concerns about vaccine safety for their pregnancy (82%, 95% CI: 78%-85%). Other reasons included concerns about vaccine safety for themselves (68%, 95% CI: 63%-72%), vaccine effectiveness (52%, 95% CI: 47%-56%), and the belief that they did not need the vaccine (22%, 95% CI: 18%-26%).

Pregnant women’s willingness to accept a COVID-19 vaccine varied by maternal race and ethnicity as well as other baseline characteristics. Women who were non-Hispanic White were more likely to be willing to accept the vaccine than women who were non-Hispanic Black or Hispanic (Figure 2). For example, 63% (95% CI: 58%-69%) of women who were non-Hispanic White stated that they were either very likely or somewhat likely to accept the vaccine whereas only 31% (95% CI: 25%-38%) of non-Hispanic Black and 35% (95% CI: 29%-41%) of Hispanic women were very likely or somewhat likely to accept the vaccine. Stated otherwise, 69% (95% CI: 62%-75%) of non-Hispanic Black and 65% (95% CI: 59%-71%) of Hispanic women were not likely to receive the vaccine compared to only 37% (95% CI: 31%-42%) of non-Hispanic White women. In addition to maternal race and ethnicity, Utah study site, having a graduate school degree, and getting an influenza vaccine during the 2019-2020 influenza season were also associated with COVID-19 vaccine acceptability (Table 2). However, in adjusted models that included race and ethnicity, educational level, and getting an influenza vaccine during the 2019-2020 influenza season, only race and ethnicity and prior influenza vaccine acceptance were significantly associated with willingness to get a COVID-19 vaccine (Table 2). Women who self-identified as non-Hispanic Black and women who identified as Hispanic had lower odds of accepting a COVID-19 vaccine compared to non-Hispanic White women (adjusted odds ratio [aOR] 0.4, 95% CI 0.2– 0.6 for each comparison). In contrast, women who reported getting the 2019-2020 influenza vaccine had higher odds of accepting a COVID-19 vaccine compare to those who did not (aOR 2.1, 95% CI 1.5-3.0). Model findings were consistent among women who completed the enrollment survey during August 9-November 14, 2020 versus November 14-December 10, 2020.

**Figure 2.**
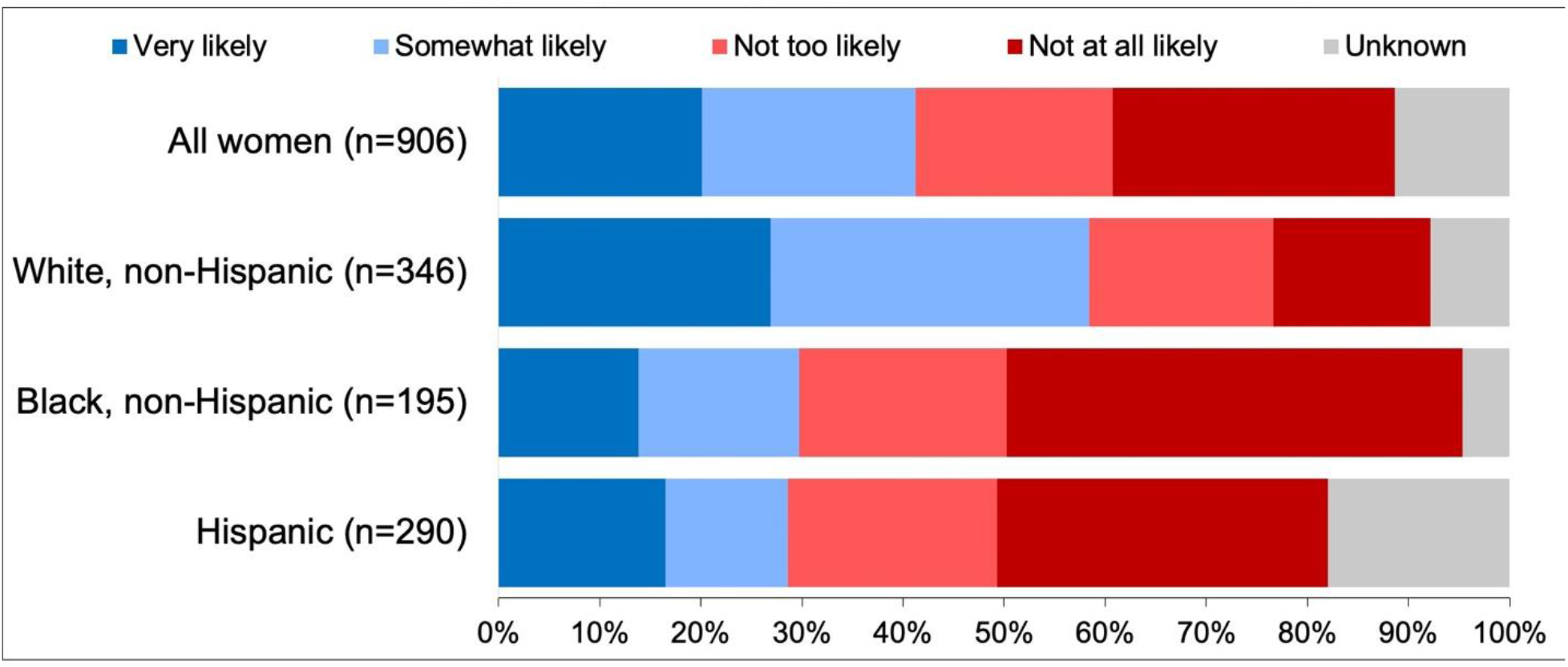
**Pregnant women’s attitudes towards SARS-CoV2 vaccination during pregnancy by race and ethnicity, N=906** Footnote: Women whose self-reported race/ethnicity was Other, non-Hispanic (n=38) and women with unknown race/ethnicity (n=37) are not shown in responses stratified by race and ethnicity.

**Table 2.** Logistic regression models evaluating factors associated with COVID-19

| Table 2. Logistic regression models evaluating factors associated with COVID-19 vaccine acceptance (n=803) |  |  |
| --- | --- | --- |
|  | **OR (95% CI)** | **aOR (95% CI)<sup>†</sup>** |
| **Site** |  |  |
| Birmingham, AL | 1.4 (1.0-2.0) |  |
| Salt Lake City, UT | **3.6 (2.5-5.1)** |  |
| New York City, NY | Ref |  |
| **Age, years (n=802)** |  |  |
| 18-34 | 0.8 (0.6-1.1) |  |
| 35-50 | Ref |  |
| **Race/ethnicity (n=773)** |  |  |
| White, non-Hispanic | Ref | Ref |
| Black, non-Hispanic | **0.3 (0.2-0.4)** | **0.4 (0.2-0.6)** |
| Hispanic | **0.3 (0.2-0.4)** | **0.4 (0.2-0.6)** |
| Other, non-Hispanic | 0.7 (0.3-1.4) | 0.8 (0.4-1.7) |
| **Education (n=802)** |  |  |
| Less than high school diploma | Ref | Ref |
| High school diploma | 0.8 (0.4-1.5) | 1.1 (0.5-2.1) |
| Some college or technical school | 0.6 (0.3-1.2) | 0.6 (0.3-1.2) |
| College degree | 1.5 (0.8-2.8) | 1.0 (0.5-2.0) |
| Graduate school degree | **2.4 (1.3-4.7)** | 1.3 (0.6-2.8) |
| **Household Below Poverty Level (n=791)** |  |  |
| Yes | 0.9 (0.7-1.3) |  |
| No | Ref |  |
| **Employed (n=797)** |  |  |
| Yes | 1.2 (0.9-1.6) |  |
| No | Ref |  |
| **Health Care Occupation (n=792)** |  |  |
| Yes | 1.4 (1.0-1.9) |  |
| No<sup>††</sup> | Ref |  |
| **Underlying Medical Condition<sup>‡</sup>** |  |  |
| ≥1 | 1.0 (0.7-1.3) |  |
| None | Ref |  |
| **Received Influenza Vaccine Last Year (n=784)** |  |  |
| Yes | **2.6 (1.9-3.6)** | **2.1 (1.5-3.0)** |
| No | Ref | Ref |
| If data are missing for a variable, n included are indicated in parentheses after variable.<br>Missing/Decline to Respond/Unknown answers not included in calculations.<br>*Willing to get vaccine defined as survey response of “very likely” or “somewhat likely”.<br>Not willing to get vaccine defined as “not likely” or “not at all likely.” Women who responded “unknown” are not included in the analysis (n=103).<br>†Final adjusted model includes variables for which aOR reported above.<br>††Includes women who are not employed.<br>‡Defined as asthma, chronic lung, metabolic, hematologic, cardiovascular, renal, hepatic, neurologic, or rheumatologic disease or hypertension.<br>Abbreviations: OR, odds ratio; CI, confidence interval; aOR, adjusted odds ratio |  |  |

Women who were worried about getting sick with COVID-19 were more likely to say they would get a COVID-19 vaccine during pregnancy than women who were not worried about getting sick (49% vs. 40%, p=0.01). Women’s worry about COVID-19 illness harming themselves and their pregnancies in the setting of COVID-19 illness was not significantly associated with vaccine acceptability (p=0.24 and p=0.60, respectively).

## DISCUSSION

In this cross-sectional study of more than 900 pregnant women from three diverse centers across the U.S., three-fourths were concerned about getting sick with COVID-19 and worried that COVID-19 could harm themselves and their pregnancies. However, less than half of pregnant women in this study said they would be likely to get a COVID-19 vaccine if one were available during their pregnancy, although this was consistent with data from a national survey of the general adult population in the US from the same period(21). Pregnant Black and Hispanic women were less likely to be willing to get a COVID-19 vaccine than White women. Women who received an influenza vaccine during the previous season were more likely to be accepting of a COVID-19 vaccine. Common perceived barriers to vaccination among women who were unwilling to get a COVID-19 vaccine included concerns about vaccine safety for their pregnancies and themselves, and concerns about vaccine effectiveness. Three in four pregnant women identified healthcare professionals as their most trusted source of information about COVID-19, with 41% specifying their obstetrician/gynecologist.

Previous studies of influenza vaccination in pregnancy have identified similar perceived barriers to vaccination to those found in our study, including concerns about vaccine safety and effectiveness and similar perceived benefits including providing protection to the pregnancy(22). Healthcare provider recommendation is one of the strongest and most consistent predictors of influenza vaccination in the United States and globally(22, 23). In our study, we also found that the majority of women identified healthcare providers as their most trusted source of information about COVID-19 and prior receipt of influenza vaccine was associated with higher odds of COVID-19 vaccine acceptability. Our findings, coupled with findings related to acceptance of other currently recommended vaccines during pregnancy, indicate that obstetricians and other healthcare providers will play a critical role in counseling pregnant women about the risks of COVID-19 illness and providing information about the safety and effectiveness of COVID-19 vaccines. Currently, both ACOG and SMFM advocate that pregnant women have the option to receive COVID-19 vaccines, and that shared decision-making be utilized by each pregnant woman and her provider regarding vaccination(16, 17). CDC also provides guidance and resources for healthcare professionals to discuss vaccination with patients before and as COVID-19 vaccines become more widely available in the United States (https://www.cdc.gov/vaccines/covid-19/hcp/engaging-patients.html).

In this study, Black race and Hispanic ethnicity were associated with lower odds of COVID-19 vaccine acceptability. These findings must be considered in the context of the well-documented history of unethical medical experimentation among racial and ethnic minority populations in the United States that has engendered mistrust of medical interventions among some communities as well as racial and ethnic disparities in key social determinants of health that may influence vaccine acceptability(24). For example, two recent studies in the United States found that Black women were less likely to report receiving a healthcare provider offer or referral for influenza or Tdap vaccination and had lower rates of receipt of both vaccines during pregnancy(25, 26). In addition, several studies have now documented that SARS-CoV-2 infection rates in the United States are higher among Black and Hispanic communities and identified disparities in COVID-19 severity among pregnant women from these groups(5, 27). For example, in a recent *Morbidity and Mortality Weekly Report*, non-Hispanic Black or African American women represented 27% of COVID-19 deaths in pregnancy, but comprised only 15% of infected pregnant women(5). In the same analysis, pregnant Hispanic women experienced SARS-CoV-2 infection at a disproportionately higher rate compared to non-pregnant Hispanic women. Our findings that women who face a greater risk of harm from COVID-19 infection are less willing to accept COVID-19 vaccination highlight the need for outreach and communication strategies to address perceived barriers to vaccination among groups that may be less likely to get a COVID-19 vaccine.

Our study has several strengths. Our population was comprised of a diverse group of pregnant women from three different centers across the United States. Efforts were made to approach all women receiving prenatal care at study clinics for study participation in the main cohort to optimize representation of the study source population. The survey collected detailed information about common perceived motivators and barriers to vaccination, tailored specifically for the contexts of pregnancy and the COVID-19 pandemic.

Several limitations should be considered when interpreting study findings. While the study was designed to maximize diversity and generalizability, only women who had access to prenatal care and who consented to participate in the prospective cohort were included. Thus, our results may not be generalizable to all pregnant women. We also did not enroll a non-pregnant group, and thus we cannot comment about how these findings may differ from a non-pregnant population in the same communities. Given that this study was conducted before a COVID-19 vaccine was available in the United States, we could not assess factors associated with actual vaccine receipt and our findings are based on hypothetical acceptability of a COVID-19 vaccine. In addition, as with all surveys, our findings may be subject to social desirability bias in which participants are more likely to respond in a manner that they perceive to be socially acceptable. Vaccine acceptance as well as perceived motivators and barriers to acceptance may evolve among pregnant women as additional data about COVID-19 vaccine safety and efficacy become available. Additional studies will be needed to monitor trends in vaccine acceptance among pregnant women over time.

Widespread vaccination is the most promising strategy to end the current global pandemic, while hand-washing, social distancing, mask wearing and other key non-pharmaceutical interventions will remain important mainstays of COVID-19 prevention. The results of this study provide insight into potential racial and ethnic disparities in COVID-19 vaccine acceptability among pregnant women. When a vaccine is available to pregnant women in the United States, obstetricians and other health care providers will play a critical role in counseling pregnant women about COVID-19 illness and offering COVID-19 vaccine to pregnant women. Similar to influenza vaccination, a clear recommendation by obstetric care professionals to pregnant women to take the vaccine will likely increase COVID-19 vaccine uptake. This may be especially important for pregnant women in racial and ethnic groups who may be at greater risk for infection and severe disease and who also appear to be less likely to accept a vaccine at this time based on the findings of our study. In order to overcome these disparities, outreach programs with community collaboration may be important to customize patient education materials that improve communication and shared decision-making in order to achieve health equity in vaccination during pregnancy.

## Data Availability

Data will automatically be sequestered by site.

## Disclaimer

The findings and conclusions in this report are those of the authors and do not necessarily represent the official position of the Centers for Disease Control and Prevention/Agency for Toxic Substances and Disease Registry, US Department of Health and Human Services.

